# NOVEL MANAGEMENT OF DEPRESSION USING KETAMINE IN THE INTENSIVE CARE UNIT

**DOI:** 10.1101/2021.09.23.21262316

**Authors:** Nirmaljot Kaur, Siva Naga S. Yarrarapu, Abhishek R. Giri, Kathleen A. Rottman Pietrzak, Christan Santos, Philip E. Lowman, Shehzad Niaz, Pablo Moreno Franco, Devang K. Sanghavi

**Author notes:** **Corresponding author:** Dr. Devang K. Sanghavi, Mayo Clinic and Hospital, 4500 San Pablo Rd. S, Jacksonville, Florida., 32224. **Funding information:** No funding received.

## Abstract

**Background:** Ketamine, a dissociative anesthetic, induces remission of depression by antagonizing glutaminergic NMDA receptors. Ketamine has been used previously in outpatient setting for treatment-resistant depression, but we showcase its utility in depression management at the Intensive Care Unit (ICU).

**Research Question:** Can ketamine be used for depression treatment in ICU patients?

**Study Design and Methods:** A retrospective chart review of ICU patients was done at a tertiary center from 2018 to 2021, to assess the ketamine usage. Among the patients reviewed, ketamine was used for depression in 12, and for analgesia & sedation in 2322 patients. Ketamine was administered in doses of 0.5mg/kg & 0.75mg/kg for depression. Each course consisted of 3 doses of ketamine administered over 3 days. 7 in 12 patients received a single course of ketamine. The rest received 3-4 courses 1 week apart.

**Results:** Ketamine was found to improve mood and affect in most of the patients with depression. 11 in 12 patients had a positive response with better sleep. It has a major advantage over conventional anti-depressants since it takes only a few hours to induce remission. Patients who refused to eat and follow treatment protocol, and who would otherwise succumb to death if not for the rapid remission of depression, were administered ketamine.

**Conclusion:** A major drawback of ketamine is that the duration of remission is short, with the response lasting only up to 7 days after a single dose. Hence, all the patients in our study were weaned off ketamine with a supporting antidepressant. Ketamine has been documented to cause cardio-neurotoxicity; however, only one patient had worsening lethargy in our study. To conclude, ketamine has a monumental benefit in treating depression in the ICU. Although our study was associated with positive outcomes, there is a need for prospective studies with long-term follow-up assessments.

## Introduction

Clinicians are increasingly using the low-dose intravenous ketamine to manage both treatment-resistant major depression and bipolar depression.^1^ Ketamine is a dissociative anesthetic that is a glutaminergic NMDA receptors antagonist.^2^ Ketamine induces a rapid treatment response, and as a result it has a substantial advantage over conventional anti-depressants which can take weeks to induce response.^3^

Depression is highly prevalent among critically ill hospitalized patients.^4^ A prospective study of 21,633 patients from twenty-six ICU centers in the UK found that 40% of patients suffered from depression and presence of depression was associated with increased mortality for two years after these patients were discharged from ICU.^5^ Patients, with serious illness and significant co-morbidities, requiring long-term intensive care are more prone to suffer from depression compared to the general population.^5^ Significant comorbid depression in these patients may prove to be fatal to the patient. By affecting the patient’s decision-making ability, it can result in treatment non-compliance or they may refuse to follow the treatment protocol. Thus, ketamine with its rapid action, can quickly induce remission and may prove to be of substantial benefit to the patient.

This case series describes our experience of using ketamine to treat depression in critically ill patients receiving care in an Intensive Care Unit (ICU) setting of a tertiary care hospital.

## Study design and Methods

This retrospective chart review study has been exempted from The Mayo clinic, Jacksonville, Florida institutional review board with identity number 21-000999. This study included all critically ill patients who were admitted to an ICU between August 2018 and February 2021. Study setting was an ICU at The Mayo Clinic, Jacksonville, Florida, a tertiary care hospital that uses a comprehensive and integrated Electronic Health Record (EHR). A retrospective chart review was conducted to identify patients who received ketamine and its indications. Patients were identified for depression based on a multimodal approach:

1. Clinical criteria: Based on the assessment of the bedside team, inputs from the nursing team, OT/PT and family.
2. Corroborating Symptoms: Other symptoms which corroborated the diagnosis were identified in these patients which includes but are not limited to insomnia, history of depression, poor participation in OT/PT, withdrawn from care team and family, restricted affect, tearfulness, voiced desire not to be alive, feeling like a burden to others, not eating and other form of exaggerated guilt.
3. Ruling out Mimics: Strong consideration was made to rule out other mimics such as delirium by CAM ICU score, psychosis, and addiction.

The approach identified above used clinical criteria, corroborating symptoms and ruling out mimics to identify depression in ICU patients who were either intubated or had a tracheostomy and were unable to use the standard PHQ-9 or other scales which usually identify depression in non-ICU patients. Primary aim of the study was to determine the tolerance and safety of ketamine. We studied the effect of administering repeated sub-anesthetic doses of ketamine (0.5mg/kg and 0.75mg/kg) to the patients with depression. The effect was measured by overall clinical improvement in addition to better sleep, better participation in OT/PT, desire to go out of the hospital room and better appetite.

Each patient was administered three consecutive doses of ketamine [Ketalar] over three days. Each intravenous infusion was given over an hour. Dosage was repeated as required.

## Results

We would like to share the results of this study in the followings cases individually.

### Case 1

A female in late 60’s with end-stage liver disease (ESLD) was transferred to the ICU due to worsening respiratory status. The patient’s hospital course was complicated by respiratory failure and shock. After more than a month-long stay in the ICU, she reported symptoms of severe depression, hopelessness, anxiety, and desire to go home. She was already taking Fluoxetine 40mg once daily and Aripiprazole 2mg once daily prior to admission. ICU physicians caring for her replaced aripiprazole with Quetiapine 150mg at bedtime and continued fluoxetine. This change in medication neither improved her anxiety nor depression in 18 days. The ICU care team recommended ketamine therapy. After 3 doses of ketamine, the patient’s mentation improved, and she slept better as evidenced by self-report and clinical presentation observed by care team members. Post ketamine therapy, the patient was maintained on Fluoxetine 40mg and Quetiapine 150mg.

### Case 2 and 3

A female in early 60’s with decompensated liver cirrhosis and a male in late 60’s with end-stage liver disease were admitted to the ICU with acute hypoxemic respiratory failure. The patient 2 was on Escitalopram 20mg daily prior to admission and was given Quetiapine 25mg during her hospital stay before starting ketamine therapy. Patient 3 was not on any anti-depressants prior to admission. During a month-long stay in ICU, both patients exhibited flat affect and clinically appeared depressed. They were prescribed ketamine. Both received a total of 9 doses of ketamine. After the course was completed, patients expressed that the new medication improved their mood as well as helped them sleep better. Post ketamine therapy, patient 2 was maintained on Escitalopram 20mg & Quetiapine 12.5mg; whereas patient 3 was started and then maintained on Escitalopram 10mg.

### Case 4

A female in late 30’s with ESLD was admitted to the ICU with hepatic encephalopathy. The patient was on Mirtazapine 30mg at night, prior to ICU admission with minimal benefits. Four days after her admission to ICU, she was prescribed ketamine for depressed mood. However, it worsened her lethargy and the second dose was not given. The ICU team attempted a second ketamine infusion after some days but it neither helped with mood nor with sleep. Post Ketamine therapy, the patient was kept on Mirtazapine 30mg.

### Case 5

A male in early 60’s was admitted to the ICU with shortness of breath. His ICU course was complicated with pneumonia. Three weeks into the hospital stay, the patient complained of poor sleep at night. The patient was on Sertraline 100mg daily prior to admission and was given Quetiapine 25mg during his hospitalization before starting ketamine therapy. After the first infusion of ketamine, the patient was more alert in the morning. He was interactive but still non-verbal. The patient stated he felt “terrible” after the first dose. However, after the 2nd and 3rd infusions, the patient felt much better. Post ketamine therapy, the patient was maintained on Sertraline 100mg.

### Case 6

A female in early 20’s initially admitted to the hospital for sever auto-immune condition, was transferred to the ICU for acute-onset hypotension. She was on Paroxetine 30mg prior to admission. During the ICU stay, she received three infusions of ketamine for symptomatic depression. The patient showed marked improvement in her mood after ketamine therapy. Post ketamine therapy, the patient was maintained on Mirtazapine 15mg at night.

### Case 7

A male in early 70’s came into the ICU with acute liver failure. The patient was not on any anti-depressants prior to admission or before ketamine therapy. Less than a month after the admission, he had waxing and waning levels of alertness along with an inability to focus for long periods of time. The patient also expressed a desire to go home but was easily redirected. The patient received a total of nine ketamine infusions. Post-infusions, the patient seemed to be in good spirits and slept well overnight. Post ketamine therapy, the patient was maintained on Sertraline 50-100mg daily and Mirtazapine 7.5mg at night. Patient was also given Quetiapine 12.5-25mg which was stopped after discharge.

### Case 8

A male in early 60’s with end-stage renal disease (ESRD) was admitted to the ICU with acute hypoxemic respiratory failure from pneumonia. The patient was not on any anti-depressants prior to admission or before ketamine therapy. About four weeks after the ICU admission, the patient was agitated and uncooperative with a flat affect and depressed mood that interfered with physical examination. He even expressed to the nurse that he did not want to leave the room and wanted to die. After twelve infusions of ketamine, the patient’s mood improved. He became very pleasant, no longer refused to leave his room and started to participate in his care. He even accepted a shave that he was previously refusing. Post ketamine therapy, he was maintained on Duloxetine 20mg once daily. Patient was also given Escitalopram 10mg once daily during his ICU stay however it was stopped at the time of discharge.

### Case 9

A female in late 50’s was admitted to the ICU, after an explorative laparotomy. The patient was on Amitriptyline 150mg at bedtime prior to admission. After spending a month in the ICU, she developed mild agitation and complained of fragmented sleep. Post-ketamine-infusions, the patient had an improved affect. She was later given Duloxetine 20mg (1 dose), Olanzapine 10mg x 11 days, Quetiapine 25mg x 14 days during her hospital course and then maintained on Amitriptyline 150mg.

### Case 10

A male in early 70’s was admitted to the ICU for continued respiratory monitoring and management of acute on chronic hypercapnic respiratory failure after laparoscopic procedure. The patient was on Trazodone 50mg at bedtime prior to admission. Ten days after the ICU admission, she was showing signs of depression and thoughts of discontinuing care. The patient showed an improved mood after the ketamine protocol. Post ketamine therapy, she was maintained on Quetiapine 50mg at bedtime.

### Case 11

A female in late 70’s was admitted to the ICU, postoperatively, after an extensive hemorrhage. The patient was on Bupropion 100mg once daily in the hospital prior to ketamine administration. After spending twenty days in the ICU, she had a depressed mood and was given three doses of ketamine. After ketamine infusions, both her mental status and depression improved. Post ketamine therapy, the patient was maintained on Bupropion 100mg.

### Case 12

A male in late 70’s was admitted to the ICU with acute hypoxemic respiratory failure. The patient was on Desvenlafaxine 25mg once daily in the ICU prior to ketamine administration. After a month-long stay in the ICU, the patient expressed that he felt extremely depressed. He received a total of three ketamine doses. Post-infusions, he was more conversant, less depressed, and less tired. Post ketamine therapy, patient was maintained on Desvenlafaxine 50mg.

## Discussion

Our case-series suggests that sub-anesthetic doses of ketamine, when administered in successive infusions, can improve mood and affect in patients with depression. Although ketamine has been used in the outpatient setting for treatment-resistant depression for many years^1^, usage of ketamine in the ICU is less common.

Our experience is consistent with the existing evidence. Treatment of depression is especially difficult in the ICU since the conventional anti-depressants take time to work (2-6 weeks). In stark contrast, according to earlier research, ketamine takes only a few hours to induce an anti-depressant response, with a maximal effect materializing at 24 hours sometimes.^1^ A significant reduction in suicidal ideation was also noted.^6^

Open-label multiple-dose IV ketamine studies have found the treatment to be safe.^7^ To date, only one Cochrane review^8^ has shown a favorable outcome for placebo as compared to ketamine specifically because it caused confusion and emotional blunting in patients with major depressive disorder. According to a study by Zhu et al.^9^, adverse effects of ketamine such as neurotoxicity, cognitive dysfunction, mental status changes, cardiovascular events, and uropathic effects are usually encountered at very high doses (5*–*40 mg/kg)^10^, when administered for prolonged periods of time, and are usually relieved by stopping the drug. In our study, only one of the twelve patients experienced worsening lethargy after the first dose of ketamine. However, after skipping a day of ketamine infusion, the patient felt good and tolerated the remaining courses better.

Although studies have shown ketamine’s ability to sustain symptom remission during the acute phase, there is a paucity of evidence with regards to its efficacy and safety profile over long-term usage (beyond two weeks). The duration of symptom remission is short, with the response lasting only up to seven days after a single dose.^7^ Another drawback of ketamine use in depression treatment is that the exact dosing and duration isn’t well studied.^11^ A meta-analysis of nine randomized control trials revealed the use of 0.40-0.80mg/kg ketamine out of which a majority of the trials used 0.5mg/kg. At our center, 0.75mg/kg was infused over 1 hour in most of the patients, and it achieved similar results.^12^

The main limitation of this review is the lack of a standardized and validated depression scale for hospitalized critically ill patients; thus, inter-observer variability may exist in the perceptions of symptomatic improvement after ketamine therapy. Likewise, many patients’ inability to communicate verbally hampers the ability of observers to assess for some depressive symptoms. Additionally, the clinical data included in our study is from one center only. Hence there is a need for more evidence from multiple ICU settings in varied locations.

To conclude, further research on the effects of ketamine in depression in critically ill patients is the need of the hour. Although our study was associated with positive outcomes, there is a need for additional prospective randomized controlled trials with both short-and long-term follow-up assessments of sample sizes with adequate power to replicate our findings.

## Conclusion

To conclude, ketamine has a monumental benefit in treating depression in the ICU. A major drawback of ketamine is that the duration of remission is short, with the response lasting only up to seven days after a single dose. Hence, all the patients in our study were weaned off ketamine with a supporting antidepressant. Ketamine has been documented to cause cardio-neurotoxicity; however, only one patient had worsening lethargy in our study. Although our study was associated with positive outcomes, there is a need for prospective studies with long-term follow-up assessments.

## Data Availability

We have all data referred to in the manuscript available with us.

## Abbreviation

ICU: (Intensive Care Unit)
NMDA: (N-methyl-D-aspartate)

## Graphic Elements

**Table 1.** Comparative analysis of different classes of anti-depressants.

|  | SSRI | SNRI | TCA | MAO inhibitors | Atypical antidepressants | Ketamine |
| --- | --- | --- | --- | --- | --- | --- |
| **Mechanism of action** | Inhibit reuptake of serotonin by binding to serotonin transporter. | Inhibit reuptake of serotonin and noradrenaline by binding to serotonin transporter and noradrenaline transporter. | Inhibit reuptake of serotonin and noradrenaline by binding to serotonin transporter and noradrenaline transporter.<br>Anti H1, H2 histamine receptors.<br>Anticholinergic | Inhibit monoamine oxidase enzyme preventing degradation of serotonin and noradrenaline. | Dopamine reuptake inhibitor.<br>Serotonin receptor modulators and reuptake inhibitors.<br>Anti $\alpha 1$ adrenergic and anti H1 histamine receptor. | Uncertain<br>*N*-methyl-D-aspartate (NMDA) antagonist. |

| **Time to symptom remission** | Approx. 4-6 weeks | Approx. 3-6 weeks | Approx. 6-8 weeks | Approx. 2 weeks | Approx. 4 weeks | Within 24 hours |
| --- | --- | --- | --- | --- | --- | --- |
| **Side effects** | Headache, sweating, dry mouth, sexual dysfunction, nausea, drowsiness | Headache, dry mouth, nausea, tremors, blurred vision, sexual dysfunction, dizziness | Headache, dry mouth, tremors, drowsiness, indigestion, increased heart rate | Insomnia, muscle aches, low blood pressure, dry mouth, sexual dysfunction, dizziness | Headache, sweating, dry mouth, sexual dysfunction, vomiting, insomnia, appetite changes, indigestion | Headache, dry mouth, dizziness, increased blood pressure, increased anxiety and psychotic symptoms, dissociative effects |
| **Indications** | Depression, phobia, panic attack, eating disorder, premenstrual | Depression, melancholic depression, | Depression, enuresis, psychotic depression | Depression, atypical depression, Parkinson's disease | Depression, smoking cessation | Depression, anesthesia, analgesia, sedative |
|  | syndrome, PTSD,<br>kleptomania,<br>obsessive-<br>compulsive<br>disorder, seasonal<br>affective disorder | psychotic<br>depression |  |  |  |  |
| **Average**<br>**Cost/month** | 12\$ | 19\$ | 4\$ | 50\$ | 17\$ | 44\$ |

**Table 2.**
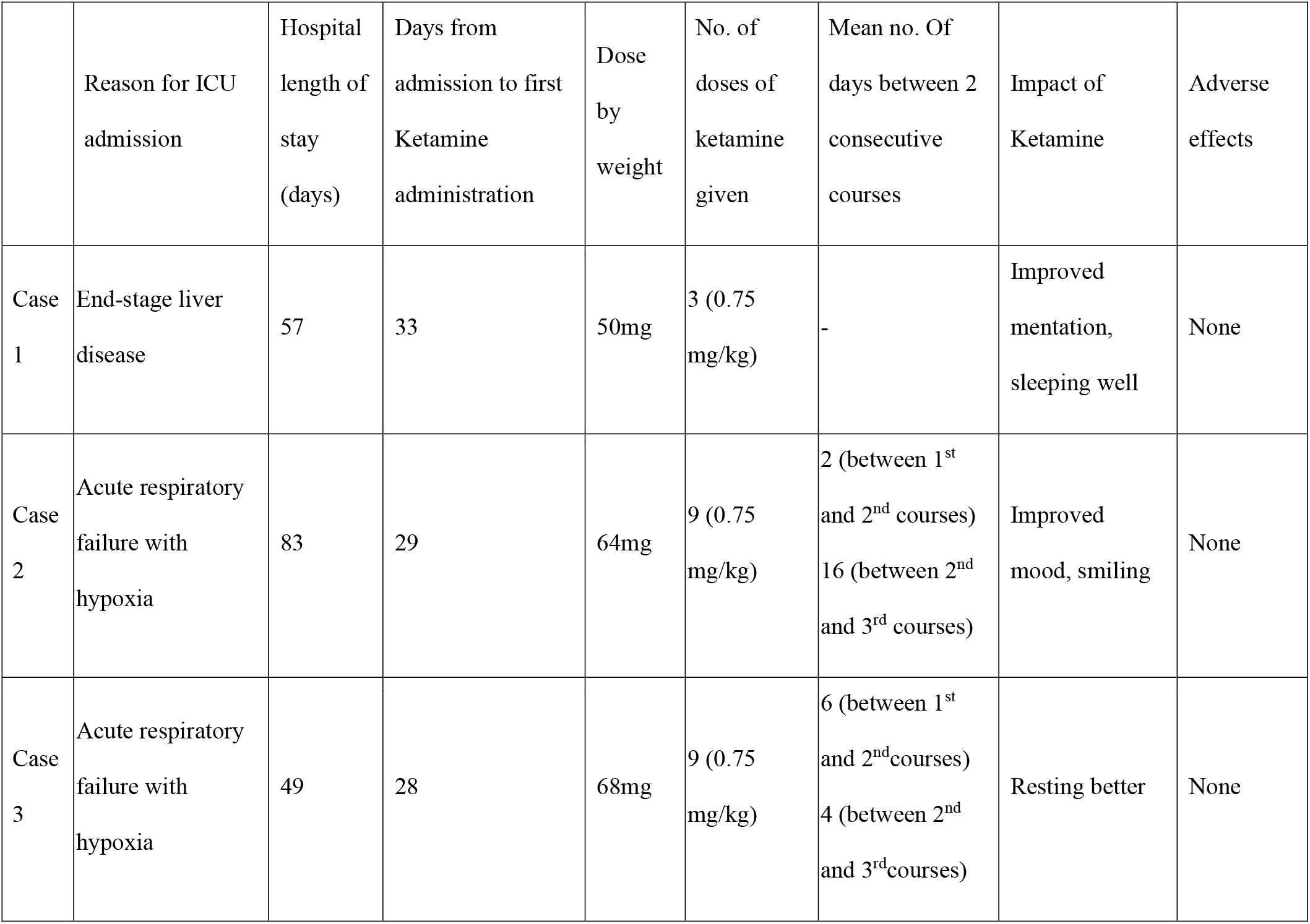
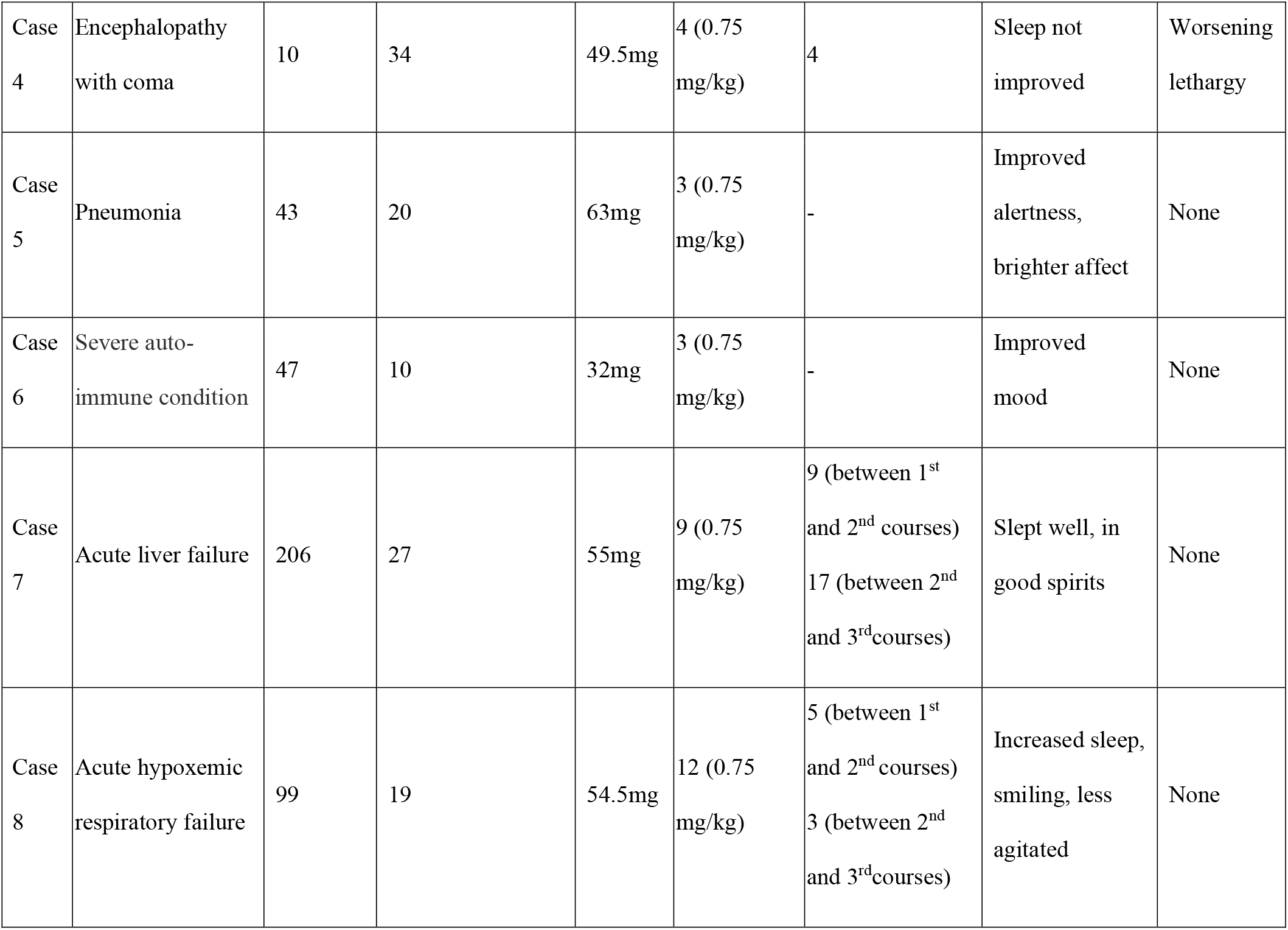
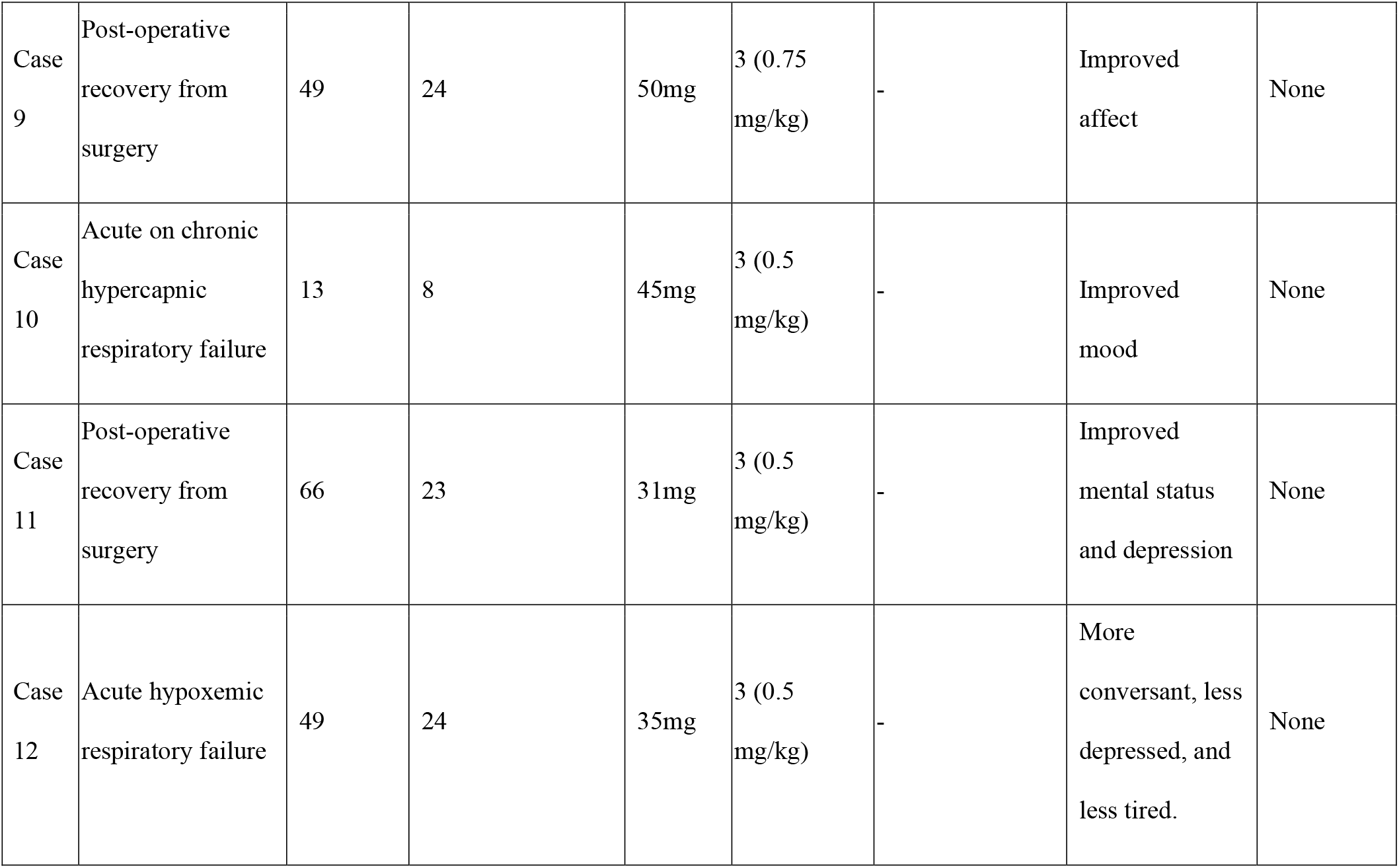
Evaluation of ketamine therapy.

|  | Reason for ICU admission | Hospital length of stay (days) | Days from admission to first Ketamine administration | Dose by weight | No. of doses of ketamine given | Mean no. Of days between 2 consecutive courses | Impact of Ketamine | Adverse effects |
| --- | --- | --- | --- | --- | --- | --- | --- | --- |
| Case 1 | End-stage liver disease | 57 | 33 | 50mg | 3 (0.75 mg/kg) | - | Improved mentation, sleeping well | None |
| Case 2 | Acute respiratory failure with hypoxia | 83 | 29 | 64mg | 9 (0.75 mg/kg) | 2 (between 1<sup>st</sup> and 2<sup>nd</sup> courses)<br>16 (between 2<sup>nd</sup> and 3<sup>rd</sup> courses) | Improved mood, smiling | None |
| Case 3 | Acute respiratory failure with hypoxia | 49 | 28 | 68mg | 9 (0.75 mg/kg) | 6 (between 1<sup>st</sup> and 2<sup>nd</sup> courses)<br>4 (between 2<sup>nd</sup> and 3<sup>rd</sup> courses) | Resting better | None |
| Case 4 | Encephalopathy with coma | 10 | 34 | 49.5mg | 4 (0.75 mg/kg) | 4 | Sleep not improved | Worsening lethargy |
| Case 5 | Pneumonia | 43 | 20 | 63mg | 3 (0.75 mg/kg) | - | Improved alertness, brighter affect | None |
| Case 6 | Severe auto-immune condition | 47 | 10 | 32mg | 3 (0.75 mg/kg) | - | Improved mood | None |
| Case 7 | Acute liver failure | 206 | 27 | 55mg | 9 (0.75 mg/kg) | 9 (between 1<sup>st</sup> and 2<sup>nd</sup> courses)<br>17 (between 2<sup>nd</sup> and 3<sup>rd</sup> courses) | Slept well, in good spirits | None |
| Case 8 | Acute hypoxemic respiratory failure | 99 | 19 | 54.5mg | 12 (0.75 mg/kg) | 5 (between 1<sup>st</sup> and 2<sup>nd</sup> courses)<br>3 (between 2<sup>nd</sup> and 3<sup>rd</sup> courses) | Increased sleep, smiling, less agitated | None |
| Case 9 | Post-operative recovery from surgery | 49 | 24 | 50mg | 3 (0.75 mg/kg) | - | Improved affect | None |
| Case 10 | Acute on chronic hypercapnic respiratory failure | 13 | 8 | 45mg | 3 (0.5 mg/kg) | - | Improved mood | None |
| Case 11 | Post-operative recovery from surgery | 66 | 23 | 31mg | 3 (0.5 mg/kg) | - | Improved mental status and depression | None |
| Case 12 | Acute hypoxemic respiratory failure | 49 | 24 | 35mg | 3 (0.5 mg/kg) | - | More conversant, less depressed, and less tired. | None |

**Table 3.**
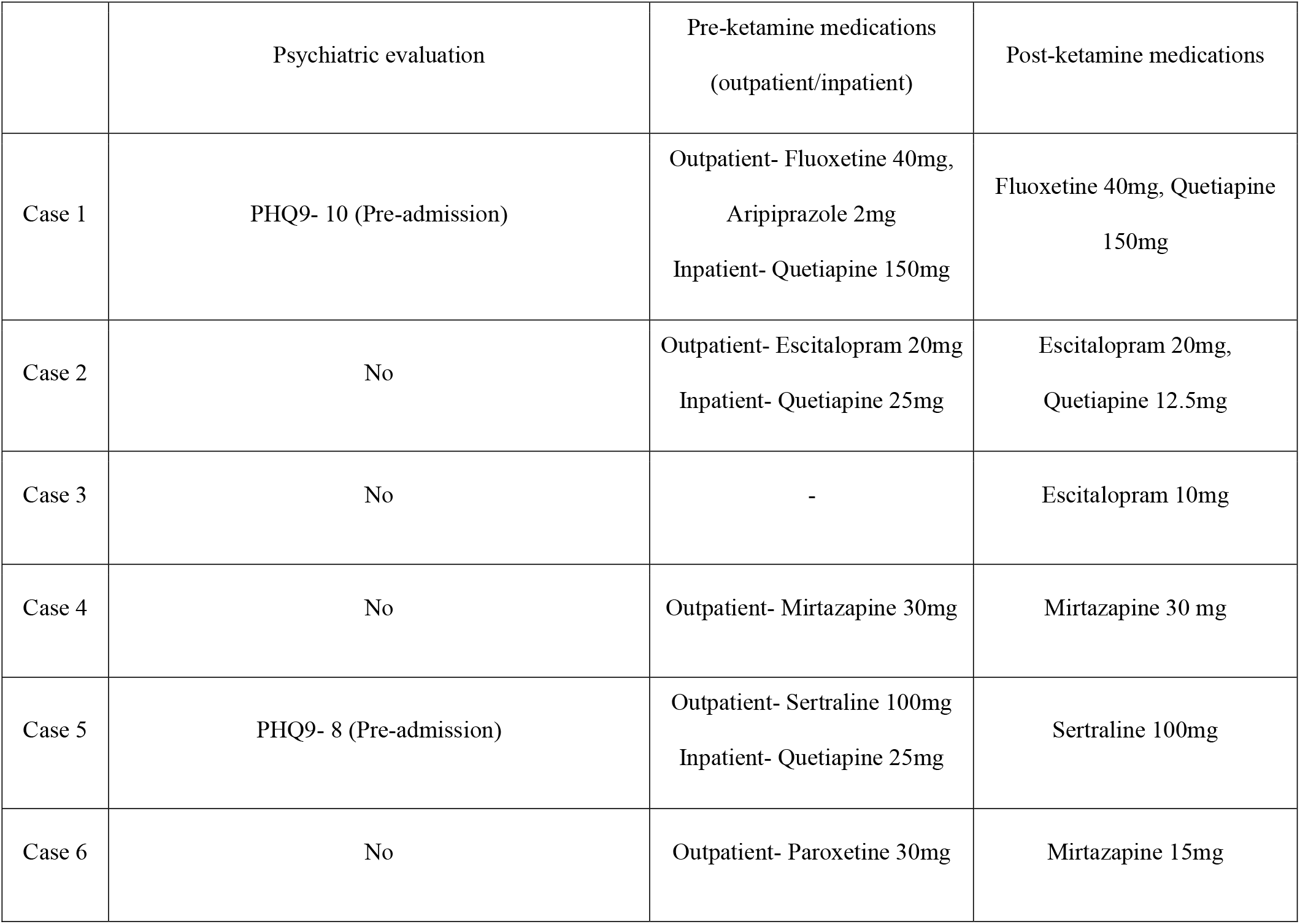
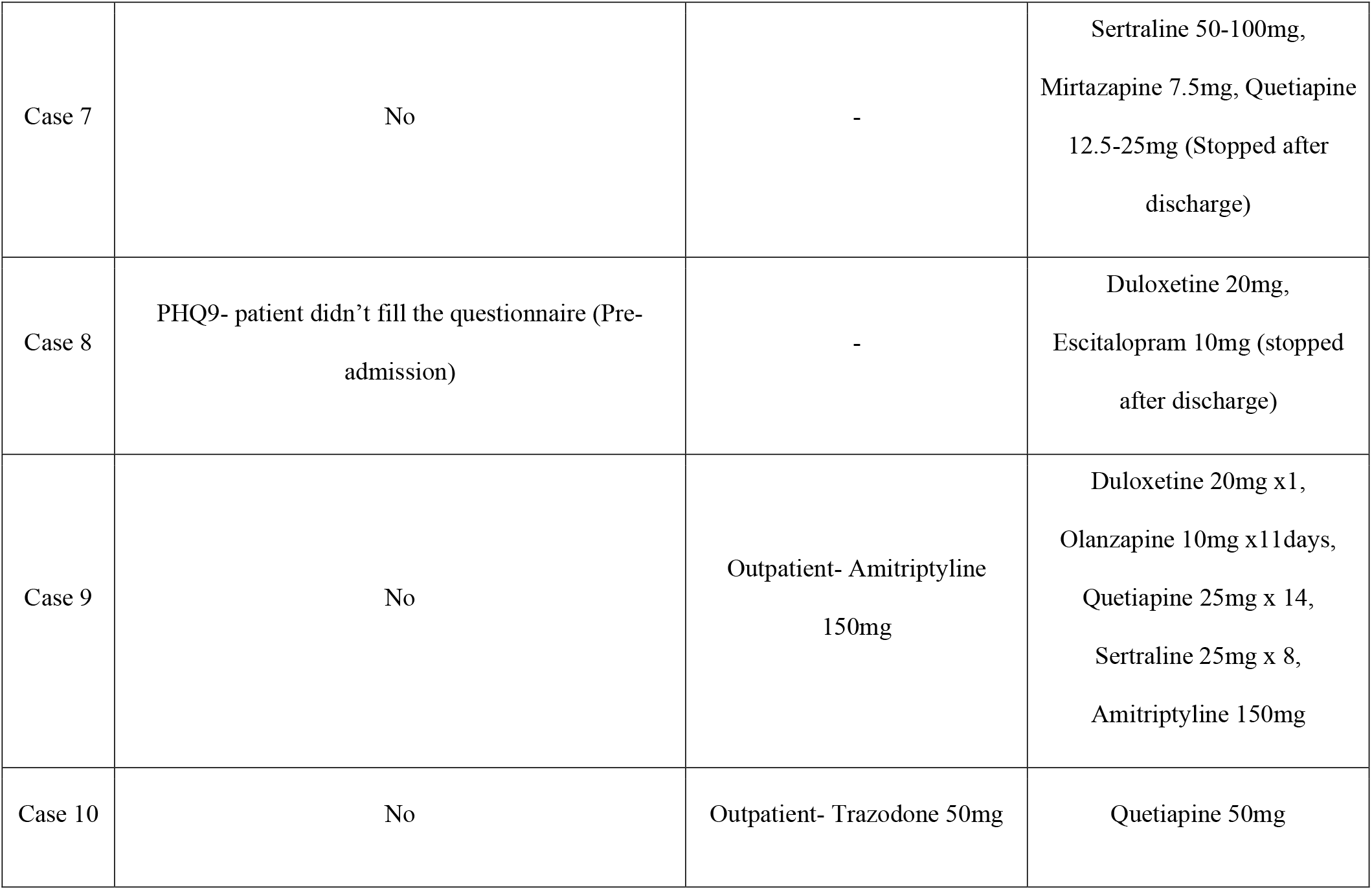
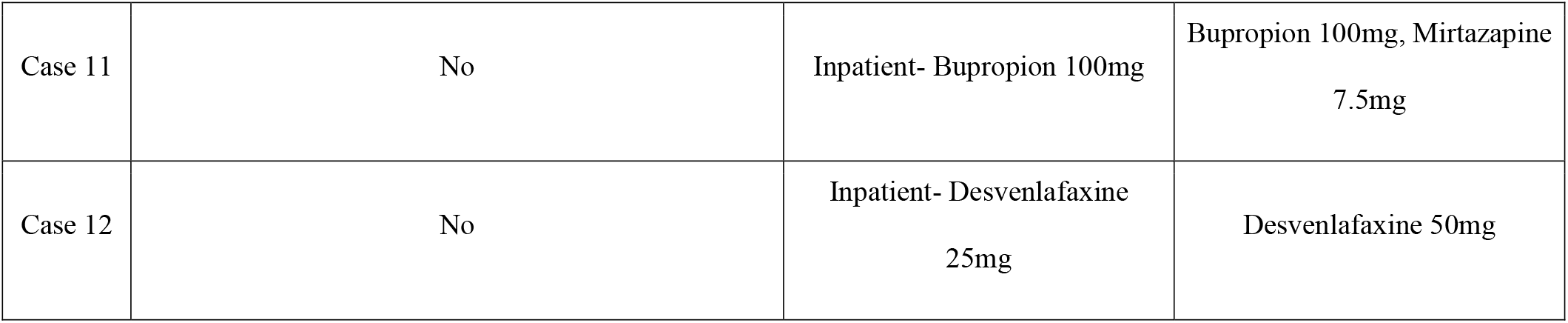
Medication administration before and after ketamine administration.

|  | Psychiatric evaluation | Pre-ketamine medications<br>(outpatient/inpatient) | Post-ketamine medications |
| --- | --- | --- | --- |
| Case 1 | PHQ9- 10 (Pre-admission) | Outpatient- Fluoxetine 40mg,<br>Aripiprazole 2mg<br>Inpatient- Quetiapine 150mg | Fluoxetine 40mg, Quetiapine<br>150mg |
| Case 2 | No | Outpatient- Escitalopram 20mg<br>Inpatient- Quetiapine 25mg | Escitalopram 20mg,<br>Quetiapine 12.5mg |
| Case 3 | No | - | Escitalopram 10mg |
| Case 4 | No | Outpatient- Mirtazapine 30mg | Mirtazapine 30 mg |
| Case 5 | PHQ9- 8 (Pre-admission) | Outpatient- Sertraline 100mg<br>Inpatient- Quetiapine 25mg | Sertraline 100mg |
| Case 6 | No | Outpatient- Paroxetine 30mg | Mirtazapine 15mg |
| Case 7 | No | - | Sertraline 50-100mg,<br>Mirtazapine 7.5mg, Quetiapine<br>12.5-25mg (Stopped after<br>discharge) |
| Case 8 | PHQ9- patient didn't fill the questionnaire (Pre-<br>admission) | - | Duloxetine 20mg,<br>Escitalopram 10mg (stopped<br>after discharge) |
| Case 9 | No | Outpatient- Amitriptyline<br>150mg | Duloxetine 20mg x1,<br>Olanzapine 10mg x11days,<br>Quetiapine 25mg x 14,<br>Sertraline 25mg x 8,<br>Amitriptyline 150mg |
| Case 10 | No | Outpatient- Trazodone 50mg | Quetiapine 50mg |
| Case 11 | No | Inpatient- Bupropion 100mg | Bupropion 100mg, Mirtazapine<br>7.5mg |
| Case 12 | No | Inpatient- Desvenlafaxine<br>25mg | Desvenlafaxine 50mg |

**Table 4.** Outcomes of ketamine therapy.

| Patients | Before Ketamine administration | After Ketamine administration |
| --- | --- | --- |
| 1 | Insomnia<br>Withdrawn from care team and family | Improved sleep<br>Improved Interaction with team and family |
| 2 | Insomnia<br>Restricted affect | Improved sleep<br>Improved expression |
| 3 | Restricted affect<br>Tearfulness | Improved Expression<br>No tearfulness |
| 4 | Insomnia | Not Improved |
| 5 | Insomnia | Improved sleep |
| 6 | Insomnia | Not Improved |
| 7 | Insomnia | Improved sleep |
| 8 | Insomnia<br>Poor participation in OT-PT.<br>Withdrawn from care team and family<br>Restricted affect<br>Voiced desire to not be alive<br>Exaggerated guilt<br>Not eating and participating in cares | Improved sleep<br>Improved participation in OT-PT<br>Improved Interaction with team and family<br>Improved Expression<br>No guilt/voiced desire to not be alive<br>Eating and participating in cares |
| 9 | Poor participation in OT-PT.<br>Withdrawn from care team and family<br>Not eating and participating in cares | Improved participation in OT-PT<br>Improved Interaction with care team and family<br>Eating and participating in cares |
| 10 | Insomnia | Insomnia not improved |
|  | Poor participation in OT-PT.<br>Withdrawn from care team and family<br>Restricted affect<br>Not eating and participating in cares | Improved Participation in OT-PT.<br>Improved Interaction with care team and family<br>Improved Expression<br>Eating and participating in cares |
| 11 | Restricted affect | Improved expression |
| 12 | Insomnia<br>Not eating and participating in cares | Insomnia not improved<br>Eating and participating in cares |

## Guarantor

Dr. Devang K. Sanghavi, M.B.B.S., M.D. takes responsibility for the content of the manuscript, including the data and analysis.

## Author contributions

All authors had full access to all the data in the study and takes responsibility for the integrity of the data and the accuracy of the data analysis, including and especially any adverse effects. Nirmaljot Kaur, Siva Naga S. Yarrarapu, Abhishek R. Giri, Kathleen A. Rottman Pietrzak, Christan Santos, Philip E. Lowman, Shehzad Niaz, Pablo Moreno Franco, Devang K. Sanghavi contributed substantially to the study design, data analysis and interpretation, and the writing of the manuscript.

